# Rapid Instructed Task Learning is impaired after stroke and associated with impairments in prepotent inhibition and processing speed

**DOI:** 10.1101/2024.03.20.24304593

**Authors:** Reut Binyamin-Netser, Anat Shkedy-Rabani, Lior Shmuelof

## Abstract

**Background:** Motor rehabilitation is a central contributor to motor recovery after stroke. This process could be hampered by stroke-associated cognitive impairments, such as the capability to rapidly follow instructions (Rapid instructed task learning, RITL). RITL was never directly studied in old adults and subjects with stroke. The aim of this study was to assess RITL following stroke and its underlying cognitive determinants.

**Methods:** 31 subjects with chronic stroke and 36 age-matched controls completed a computerized cognitive examination that included an anti-saccade task for measuring prepotent inhibition and processing speed and stimulus-response association task (NEXT) for measuring RITL and proactive inhibition.

**Results:** RITL abilities were impaired after stroke, together with prepotent inhibition and processing speed. A correlation analysis revealed that RITL is associated with prepotent inhibition abilities and with processing speed.

**Conclusions:** Subjects with stroke show impairments in the ability to follow instructions, that may be related to their impaired prepotent inhibition and processing speed. The causal effect of RITL impairments on the responsivity to rehabilitation and on motor recovery should be examined.

## Introduction

Stroke is associated with long-term cognitive impairments. One year after stroke, 31% of survivors still show cognitive impairments (Nys et al., 2005). Cognitive impairments following stroke contribute to functional impairments and are associated with lower chances of independence after discharge (McDowd et al., 2003; Mok et al., 2004; Pohjasvaara et al., 2002; Tatemichi et al., 1994) and with poor rehabilitation outcomes (Aprile et al., 2021).

An essential cognitive ability for rehabilitation is the ability to follow instructions. This ability was studied in the lab using the rapid instructed task learning (RITL) paradigm (Cole et al., 2013), which captures the ability of subjects to follow instructions without practice. This ability is likely to be unique to humans(Verrico et al., 2011), rely on proactive cognitive control (Cole et al., 2018), and associated with increased neural activation in lateral prefrontal and posterior parietal cortices (Cole et al., 2010; Hartstra et al., 2011; Ruge & Wolfensteller, 2010).

Despite the potential role of RITL abilities in rehabilitation and daily functions, it was not studied in subjects after stroke and during healthy aging. Studies regarding individual differences in RITL abilities in young adults, suggest that these differences are driven by executive functions such as working memory (WM) (Meiran et al., 2016; Pereg & Meiran, 2019).

Several reasons suggest that RITL abilities will be reduced following stroke. First, executive functions deteriorate following stroke (Ballard et al., 2003; Cumming et al., 2013; Kase et al., 1998; Levine et al., 2015; Wall et al., 2015). Specifically, the executive functions that were found to be impaired are flexibility, inhibition, shifting and fluency (Pohjasvaara et al., 2002; Skidmore et al., 2023; Zinn et al., 2007). These functions contribute to high level cognitive abilities, such as the ability to use strategies, which are important for engagement in rehabilitation and for independent functioning in society. Second, processing speed, which is important for interpreting instructions and carrying them out, is impaired after stroke (Finkel et al., 2005; Salthouse, 1994, 2005).

The aims of this study are to (1) estimate RITL impairments after stroke (2) examine the association between inhibition and processing speed to RITL abilities and (3) highlight the possible cognitive determinants of RITL impairment after stroke.

We hypothesize that subjects with stroke (SwS) will show RITL impairments compared to age-matched controls (AMC) due to their impairments in inhibition and processing speed.

## Methods

### Subjects

31 SwS (55-72 years, mean: 65.35 years SD 5.69, 9 females) at the chronic stage (mean time post stroke=11.61 months, SD=10.04) (see Table 1 for demographic details for the stroke group - contact the corresponding author) and 36 AMC subjects (55-75 years, mean age: 67.17, SD 5.38 years, 22 females) participated in the study. SwS were recruited at Adi Negev Nahalat Eran rehabilitation center (convenience sample). AMC subjects were recruited at Ben-Gurion University of the Negev. The study was approved by the Ben-Gurion institutional Helsinki committee (for AMC subjects) and by the Regional Ethical Review Board at Sheba Medical Center, Israel (Approval Number 6218-19-SMC) (for SwS). All subjects signed a consent form before performing the experiments and were paid for their participation. SwS had a motor and sensory evaluation (motor and sensory upper extremity Fugl-Meyer assessment (FMA) and the Action Research Arm Test (ARAT)) and went through an examination of eye movements (saccades and pursuit tests) before the clinical tests. SwS did all the tasks with their unaffected arm to minimize the effect of motor impairment on task’s performance. The unaffected arm received a full motor and sensory score in the FMA and ARAT assessment in all the SwS (FMA and ARAT from both arms were obtained as part of a bigger longitudinal project). AMC subjects were instructed to use their dominant or non-dominant arm to control for hand dominance in the stroke group (22/31 stroke and 18/36 AMC subjects performed the task with their non-dominant arm).

Inclusion criteria: Ability to give informed consent and understand the tasks involved. Ages: 55-75. SwS with a first-ever stroke (ischemic or hemorrhagic) confirmed by CT or MRI or with a recurrent stroke, if their cognitive abilities and motor control were normal before the incidence (based on their medical history). Time after stroke onset ≥ 6 months. Normal eye movement and an intact ability to perceive images on the screen.

Exclusion criteria (All groups): history of physical or neurological condition that interferes with study procedures or assessment of motor function (e.g., severe arthritis, severe neuropathy, Parkinson’s disease). SwS that were dependent before the incident with documented motor or cognitive impairments. Subjects that did not complete 75% of the cognitive battery were also excluded.

All data produced in the present study are available upon reasonable request to the authors.

### Experimental design

The study was composed of computerized cognitive examination and a clinical cognitive assessment. The computerized cognitive examination was composed of two tasks: (1) Anti-saccade, (2) NEXT paradigm. The clinical cognitive assessment was the Montreal Cognitive Assessment (MoCA) (Nasreddine et al., 2005), a one-page assessment that tests multiple cognitive domains: visuospatial perception (5 points), naming (3 points), attention (6 pints), language (3 points), abstraction (2 points), short-term memory (5 points) and orientation (6 points). The maximal score in MoCA is 30 points. This test is a clinical cognitive screening tool that shows high sensitivity and specificity in identifying poststroke cognitive impairment in both ischemic and hemorrhagic strokes (Chiti & Pantoni, 2014). As part of the intake, SwS went through motor and sensory evaluation (FMA and ARAT) and also went through an eye movement test (saccades and pursuit manual tests).

## Computerized cognitive battery

**(1) Anti-saccade task:** This task is used for examining prepotent inhibition abilities (Katzir et al., 2010; Miyake et al., 2000; Roberts et al., 1994). At the beginning of the task, a fixation point is presented in the middle of the screen for a variable duration (1500-3500 ms). Then, a visual cue (a white circle shaped symbol) is presented on one side of the screen (right or left 10.6 cm from the center) followed by a target stimulus (a white arrow in a square) that is presented in the opposite side to the visual cue. The target stimulus appears after a delay of 200, 300, 400 or 500ms. The delay for each trial was randomly selected. The target stimulus was presented for 166 ms and then masked by a white square. The subject’s goal was to respond according to the direction of the arrow in the target stimulus (up, down, let, right) using the arrow keys of the keyboard. Subjects had 2000 ms to respond. If they failed to respond, the trial ended with an error and the next trial began. Subjects received feedback after each trial. The feedback was a green V shape with a cheerful sound for correct answers or a red X shape with buzz sound for incorrect answers, presented on the center of the screen. Subjects were instructed to respond as accurately as they can. The task consisted of 3 blocks (40 trials each) separated by a break. To increase the difficulty of the task, the visual cue did not disappear when the target stimulus appeared. Because the target stimulus appears at a random delay and for a very short time, subjects had to inhibit their automatic saccade to the first appearing visual cues to succeed in identifying the direction of the arrow and answer correctly. The primary variable of interest is accuracy (percentage of correct answers) which reflects prepotent inhibition abilities (Figure 1.A).
**(2) The NEXT paradigm:** This paradigm (Meiran et al., 2015) was designed to quantify the ability to follow instructions - known as rapid instructed task learning (RITL) (for review see (Cole et al., 2013)), and proactive inhibition - the ability to ignore information that is irrelevant to the instructed task (Friedman & Miyake, 2004). In every mini-block, two kinds of trials are presented: GO (2 trails) and NEXT trails (0-5 trails). NEXT stimuli appear in red and GO stimuli appear in green. In the instruction screen, subjects are introduced with two stimuli (symbols or pictures) one on the right and the other on the left sides of the screen, which indicate their association with two keyboard keys (left and right arrow keys). Then, one of the two prior presented stimuli appears in the middle of the screen and the subject had to press the instructed key. In the NEXT phase, subjects were instructed to respond with the right or left arrow keys, throughout the experiment, based on a global instruction that they received at the beginning of the experiment. In the GO phase, subjects were asked to respond according to the mapped stimuli (right or left key) that were changed between mini-blocks.

Each mini-block began with an instruction screen, continued with NEXT trails and ended with GO trails. Before each trial, a fixation point appeared for 500ms (with a jitter that is drawn from Gaussian distribution with a standard deviation of 50 ms). To keep subjects always ready to perform the GO task, the number of NEXT trials vary between mini-blocks (in every block there are 10% of zero, 30% of one, 20% of two, 20% of three, 10% of four and 10% of five NEXT trails). The GO phase consists of only two trials per mini-block. These constraints forced subjects to be alerted. There was no time limit for pressing the answer keys; Each stimulus was presented until the subject pressed the answer key. Trials were separated by 500ms. At the end of each mini-block, subjects received feedback regarding the accuracy and the average RT of the two GO trials that were presented at the end of the mini-block. The feedback was presented for 3 seconds and was followed by the next mini-block. Subjects were instructed to try to be as fast and as accurate as they can. The paradigm consisted of a training phase (that made sure subjects understood the task) and 60 mini-blocks (separated to 6 blocks). Each mini-block displayed new stimuli.

**Figure 1:**
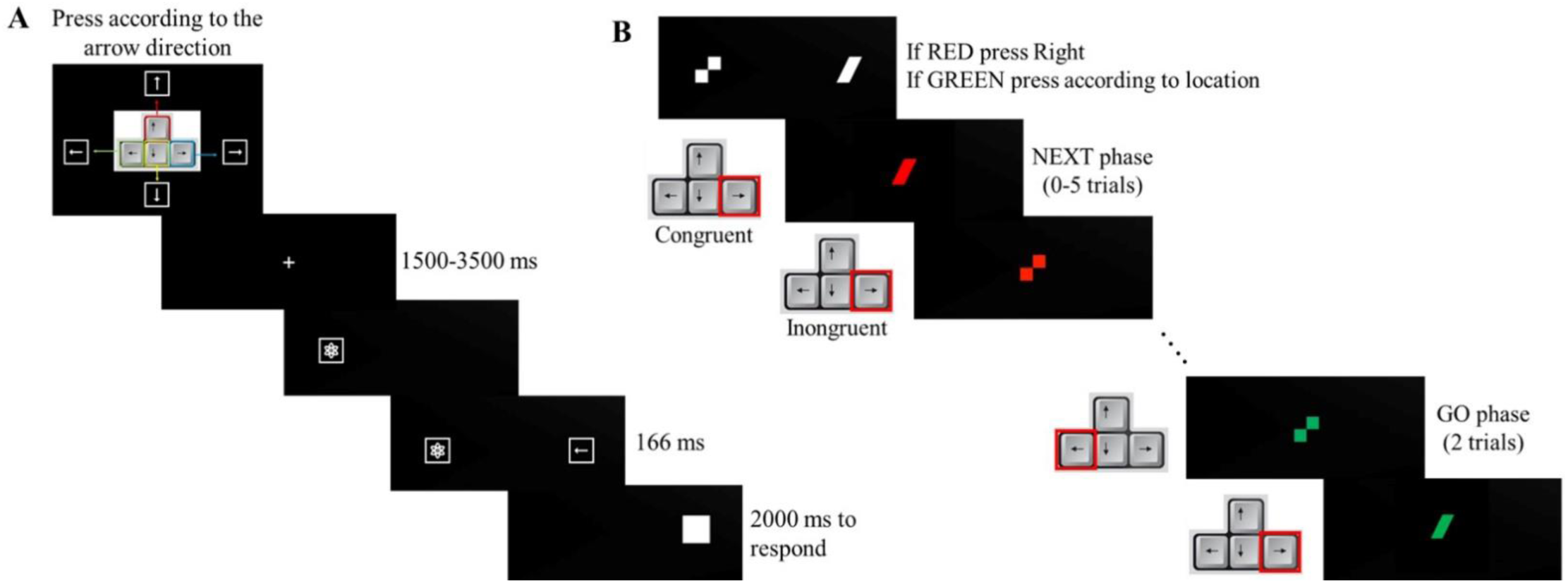
**A.** The Anti-saccade task. **B.** The NEXT paradigm.

There was one break in the middle of the task. Two effects of interest were examined: (1) *Proactive inhibition - The NEXT effect*: the faster reaction time when the NEXT response key was congruent with the GO mapped key (for example, the NEXT key is the left arrow key and the appeared red stimulus was mapped to the left arrow key) compared to the opposite (incongruent) condition (the NEXT key is the left arrow key and the appeared red stimulus was mapped to the right arrow key). (2) *Rapid instructed task learning (RITL)*: accuracy in the first (GO1) and second (GO2) trials and enhancement of response between the first and second GO trials (GO effect). Smaller GO effect suggests that the subject is more efficient in the GO1 response (Figure 1.B). importantly, no feedback was provided after the GO1 response.

### Preregistration

This study is a part of a larger study that was preregistered online https://osf.io/2y5jx.

### Power analysis

Power was computed based on the anti-saccade task, where effect sizes were reported (Arbiv & Meiran, 2015; Hull et al., 2008). Effect size was set to 0.8 based on data from healthy subjects. With alpha of 0.05, and power of 0.8, the estimated number of subjects was 21 per group for comparison of means of two groups.

### Data analysis

The computerized tasks were performed using OpenSesame version 3.2.7. All analyses and statistical calculations were performed in MATLAB 2020a (The MathWorks) and SPSS 21 (IBM SPSS statistics). In the anti-saccade task, RT was limited to 2000 ms. In the NEXT paradigm, where RT was not limited experimentally, RTs lower than 150 ms or higher than 4000 ms were trimmed. The average RT was calculated only from the correct response trials in all tasks.

All the SwS that were enrolled in the study were after a first stroke (no recurrent strokes in the sample). All subjects were included in the final analyses except one stroke subject that was excluded due to incomplete data (less than 75% of collected data). Data of the Anti-saccade task of one subject with stroke was excluded due to a ptosis of the eyelid. Data of the NEXT task of one subject with stroke was also excluded since he didn’t understand the task.

### Statistical analysis

#### RITL analysis

RITL was assessed based on the accuracy (error rate) in GO1 and GO2 and based on the GO effect – the RT enhancement between GO1 and GO2. Statistical comparison between the two groups (AMC or SwS) was performed using an independent sample t-test. Z scores were obtained for each parameter separately for each group. A subject with a z score bigger than ±3.3 was removed from the analysis.

#### Prepotent inhibition analysis

Prepotent inhibition is the ability to ignore distractors. It was calculated by computing the proportion of correct responses in the anti-saccade task. Statistical comparison between the two groups (AMC or SwS) was performed using an independent sample t-test. Z scores were obtained for each parameter separately for each group. A subject with a z score bigger than ±3.3 was removed from the analysis.

#### Proactive inhibition analysis

Proactive inhibition, is the ability to ignore information that is irrelevant to the instructed task. It was represented by the NEXT effect which is calculated by subtracting the RTs in the congruent NEXT trials from the RTs in the incongruent NEXT trials. Only the first two NEXT trials were taken from each mini-block. NEXT was computed only in cases where the GO trials in the same mini-block were answered correctly, to make sure subjects remembered the association. Statistical comparison between the two groups (AMC or SwS) was performed using an independent sample t-test. Z scores were obtained for each parameter separately for each group. A subject with a z score bigger than ±3.3 was removed from the analysis.

#### Processing speed analysis

Processing speed was estimated based on the RT in the anti-saccade paradigm. The RTs were taken only from the correct trials. Statistical comparison between the two groups (AMC or SwS) was performed using an independent sample t-test. Z scores were obtained for each parameter separately for each group. A subject with a z score bigger than ±3.3 was removed from the analysis.

#### Correlation analyses

Correlation analyses were made between RITL and the cognitive parameters using Pearson correlation test. Before each correlation, observations that deviate by more than 3.3 SDs from the group’s distribution (outliers) were removed. Outlier’s removal can be seen through the degrees of freedom in the correlation analyses.

#### Linear regression analysis

Linear regression between RITL and the sub-cognitive domains of MoCA was conducted using a regression model where RITL was the dependent variable and the score of the sub domains - perception, naming, attention, language, abstraction and short-term (ST) memory were the independent variables. The sub-cognitive domain – orientation did not enter the model because all subjects received a full score in this domain. Reported betas are standardized. Confidence interval was 95%.

Multicollinearity was examined for the regression model that was performed. No independent variables were highly correlated with each other (r>0.8). In order to find influential outliers, a Cook’s distance test was applied before performing all the regression analyses. No outliers were found (Di<1).

## Results

Multiple variables in our design are associated with the ability of subjects to rapidly learn from instructions (RITL abilities). Primarily, the accuracy at the first trials of the 2- choice stimulus response association task (GO1 and GO2 trials) and the enhancement in response between the first and second trials (readiness potential – GO effect (Meiran et al., 2016)). All these measures were affected by stroke (GO1 error rate, t(62)=-3.08, p=0.003, *d*=0.75; GO2 error rate, t(63)=-2.99, p=0.004, *d*=0.57; GO effect, t(63)=-2.70, p=0.009, *d*=0.66 (Figure 2A & 2B)). To limit the number of comparisons, we chose to focus on the error rate in the GO2 trial as a measure of RITL. We chose the GO2 trial since it is less susceptible to switching mistakes from the prior NEXT task than the GO1 trial, and since it is not affected by learning as no feedback was provided after GO1.

**Figure 2:**
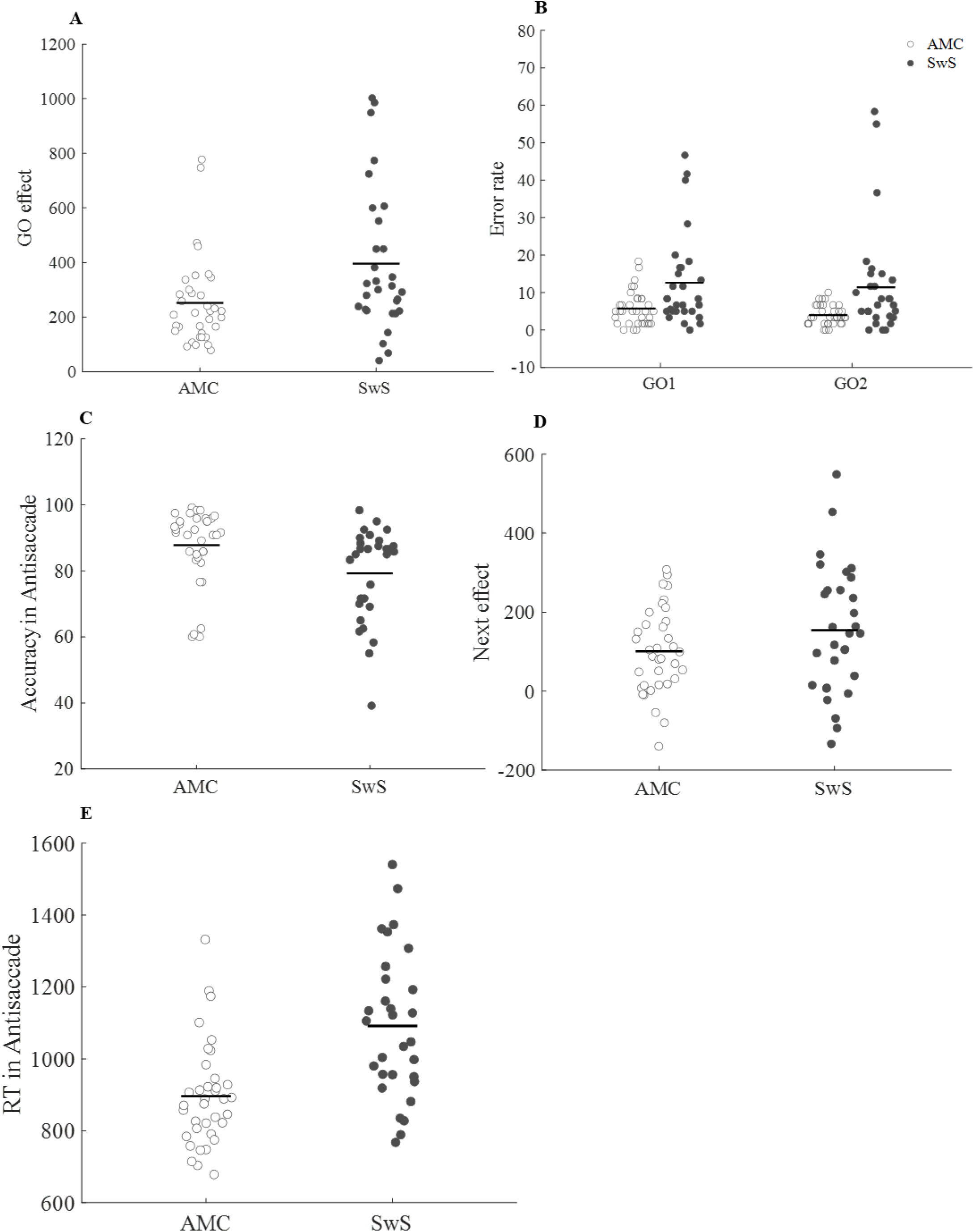
**A.** GO effect. The effect is presented for the AMC (empty dots) and SwS (filled dots) groups. **B.** Error rate in the first and second GO trials from the NEXT paradigm, divided to groups – AMC (empty dots) and SwS (filled dots). **C.** Prepotent inhibition: Average of accuracy rates in the anti-saccade task in the AMC group (empty dots) and SwS group (filled dots). **D.** Average RT in the NEXT paradigm, in the AMC (empty dots) and the SwS group (filled dots). **E.** Processing speed, average of RT in the anti-saccade task for the AMC group (empty dots) and SwS group (filled dots).

Next, we searched for the possible underlying cognitive determinants of this ability. Two types of inhibition were examined: (1) prepotent inhibition and (2) proactive inhibition. Prepotent inhibition was examined using the accuracy in the anti-saccade task. Proactive inhibition was examined based on the interference effect of the GO task on the NEXT task (see Methods), and was measured as the difference in RT between the NEXT congruent and incongruent responses, as measured in the NEXT task (NEXT effect). Another process that could contribute to RITL abilities is processing speed. Processing speed was estimated based on the RT in the anti-saccade paradigm in successful trials. A significant association was found between RITL ability and the three examined cognitive components: prepotent inhibition (r(62)=-0.26, p=0.04; Figure 3A), proactive inhibition (r(62)=0.33, p=0.008; Figure 3B) and processing speed (r(62)=0.27, p=0.03; Figure 3C).

**Figure 3:**
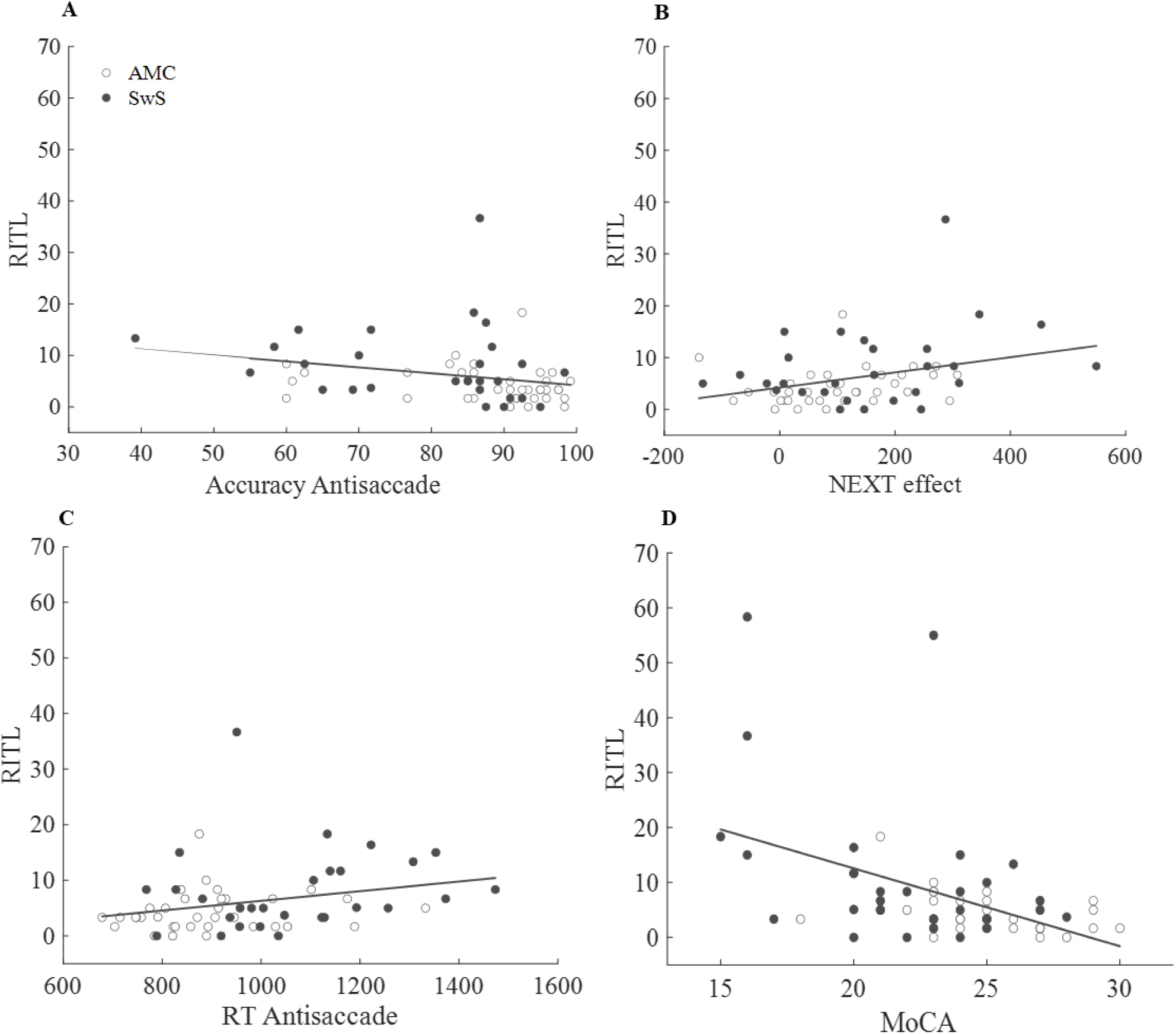
scatter plots of RITL and cognitive parameters and MoCA for the AMC (empty dots) and SwS (filled dots) subjects. The continues line is a linear trendline of the entire sample. **A.** Correlation between RITL (error in second GO trial) and prepotent inhibition **B.** Correlation between RITL and proactive inhibition. **C.** Correlation between RITL and processing speed. **D.** Correlation between RITL and MoCA.

To examine the potential contribution of each component to the stroke related RITL deficits, we tested the effect of stroke on each component. Accuracy in the anti-saccade task was significantly lower in SwS compared to the AMC subjects (t(63)=2.72, p=0.008, *d*=0.67) (one outlier observations in the SwS group was excluded from the analysis). This measure indicates that prepotent inhibition abilities of SwS are impaired (Figure 2C). For the proactive inhibition parameter, both groups showed a significant NEXT effect: the RT of the incongruent trials was significantly higher than the RT of the congruent trials (SwS, RT incongruent=1218.7ms, RT congruent=1064.5ms, t(29)=-5.24, p<0.001, *d*=0.59; AMC, RT incongruent=1005.1ms, RT congruent=904.4ms, t(35)=-5.63, p<0.001, *d*=0.50). Nevertheless, no differences between the groups were found (t(64)=-1.64, p=0.11, *d*=0.39) suggesting that proactive inhibition cannot explain the RITL deficits of the stroke group (Figure 2D). For processing speed, we found that SwS had an overall slower RTs than AMC subjects (SwS 1091.99 ms (SD 203.45), AMC 896.40 ms (SD 143.32); t(64)=-4.57, p<0.001, *d*=1.11) (Figure 2E). This corresponds with other studies that show reduced processing speed after stroke (Ballard et al., 2003; Su et al., 2015). These results indicate that prepotent inhibition and processing speed impairments may underlie the RITL impairments in the SwS group.

Last, we examined the association between RITL and general cognitive assessments (MoCA), which was by itself sensitive to stroke (SwS: MoCA =21.97 (SD 3.66); AMC: MoCA =24.89 (SD 2.58); (t(65)=3.81, p<0.001, d=0.92)). A significant correlation between RITL abilities and MoCA was found (r(62)=0.51, p<0.001) (Figure 3D) indicating impairments in general cognitive abilities are associated with RITL impairment. Next, we explored the association of sub-cognitive domains (from MoCA) with RITL using a linear regression analysis. We found that attention (β=-0.29, t=-2.19, p=0.03) and short-term memory (β=-0.28, t=-1.98, p=0.05) contributed significantly to the RITL measure (Table 3). Due to the number of independent variables, we did not run this model with an interaction term with group.

**Table 3:** Multiple regression analysis of sub cognitive domains of MoCA.

|  | beta | t | p |
| --- | --- | --- | --- |
| **Perception** | 0.06 | 0.41 | 0.69 |
| **Naming** | -0.20 | -1.78 | 0.08 |
| **Attention** | -0.29 | -2.19 | **0.03\*** |
| **Language** | -0.09 | -0.70 | 0.49 |
| **Abstraction** | -0.02 | -0.19 | 0.85 |
| **ST memory** | -0.28 | -1.98 | **0.05\*** |

## Discussion

Our results show that RITL abilities are decreased after stroke. In search for the cognitive determinants of this ability, we report that prepotent inhibition, proactive inhibition and processing speed are associated with RITL abilities. Furthermore, we found that processing speed and prepotent inhibition abilities are reduced due to stroke while proactive inhibition abilities are not. In our exploratory quest we also found an association between RITL and general cognitive measures of attention and short-term memory.

### RITL impairment in stroke

Stroke is associated with multiple cognitive impairments (Ballard et al., 2003; Cumming et al., 2013; Kase et al., 1998; Levine et al., 2015; Tatemichi et al., 1994; Wall et al., 2015). Mild cognitive impairments may not be detected when high cognitive functions such as learning and the use of strategies are examined (Binyamin-Netser et al., 2023) probably since subjects after stroke use additional general cognitive resources (Goh & Park, 2009). In this case, we detected the cognitive impairment in RITL, which may be considered a high cognitive function. One explanation for the detected impairment in such as task could be that the cognitive load that the task demanded was too high and SwS did not have enough cognitive resources to compensate for the impairment. Another explanation could be that some of the cognitive determinants of RITL, such as processing speed and inhibition, (Chow et al., 2020), cannot be compensated for. We speculate that by repeating the same stimuli for several trials, with feedback after each trial, will bring all subjects to a high level of performance, and could allow for subjects with cognitive impairment to compensate for their RITL impairments.

### Underlying components of RITL

RITL ability was previously shown to depend on working memory processes in young adults (Bergman-Nutley & Klingberg, 2014; Dunham et al., 2020; Meiran et al., 2016; Pereg & Meiran, 2019). We report here that inhibition (prepotent and proactive) and processing speed are also an underlying component of RITL in old adults and SwS. This dependence may be specific to these groups, that are known to use more general cognitive resources (Boisgontier et al., 2013; Cumming et al., 2013; Heuninckx et al., 2008; Hom & Reitan, 1990; Tatemichi et al., 1994; Wall et al., 2015) or represent a general feature of RITL. Furthermore, while inhibition was not studied in the context of RITL, it was shown that the ability to follow instruction is influenced by metacognition (Dunham et al., 2020). We therefore suggest that inhibition, a central component in executive function (Roebers & Feurer, 2016), contributes to RITL also in young adults, and call for expanding the effort to study the underlying cognitive determinants of RITL in young adults.

### Prepotent and proactive inhibition

Inhibition abilities have been studied in adults compared to young participants. The results have shown that prepotent inhibition abilities are declined with age in comparison to proactive inhibition which remains intact (Arbiv & Meiran, 2015; Hull et al., 2008; Katzir et al., 2010; Rey-Mermet et al., 2018). There are only few studies regarding inhibition abilities after stroke. Only one of them investigated oculomotor inhibition (Peterburs et al., 2011) and showed that stroke subjects had more errors and had longer saccade latencies. Other studies reported that inhibition abilities are reduced after stroke (Ballard et al., 2003; Laakso et al., 2019; Pohjasvaara et al., 2002; Zinn et al., 2007) but did not differentiate between prepotent and proactive inhibition. The qualitative consistence of the results of the SwS with the results in old adults (Rey-Mermet et al., 2018) supports the notion that stroke cause an enhancement in brain atrophy (Brodtmann et al., 2020; Graham & Sharp, 2019), particularly in the frontal lobes (dorsolateral prefrontal cortex) that are essential for executive functions (Possin et al., 2014; Stuss & Levine, 2002).

### Implications for rehabilitation

Our results indicate that RITL abilities are impaired after stroke thereby suggesting that compliance during rehabilitation may be affected by the cognitive decline of the subjects. Thus, to improve rehabilitation compliance and outcomes, RITL abilities should be monitored, and clinicians should consider using simpler instructions and repeating the instructions from session to session. In the context of self-training and tele-rehab, specific emphasis should be given to the instructions and the initial compliance of the subjects should be analyzed with respect to the ability to understand the instructions. The finding that inhibition (prepotent and proactive) and processing speed may underlie the RITL impairments suggests that in order to minimize the impact of the RITL impairment on rehabilitation, therapy sessions should take place in a quiet place with minimum stimulations and should concentrate on one instruction at a time with substantial response times.

### Limitations

One limitation is the assumption that the cognitive measures that we investigate represent distinct cognitive abilities (Dubois et al., 2018); and that integral cognitive components (such as working memory) are missing from our design. We therefore call for replicating our results and for investigating the effect of parametrically manipulating the load of inhibition, processing speed, and working memory, to better assess their differential contribution to RITL and to examine the role of additional cognitive abilities in RITL. An additional limitation is the small sample size that was powered for between group comparisons but not for running regression models. We suggest that this matter should be addressed in a larger sample study, or in a study that focus only on specific interactions. Last, this study focused on chronic subjects after stroke with different cognitive impairment levels. It could be that subjects with lesions to specific neural substrates or with sever cognitive impairments will show additional cognitive impairments and RITL determinants.

## Conclusion

The ability of subjects after stroke to follow instructions rapidly is impaired and associated with their impairments in pre-potent and proactive inhibition and processing speed. Cognitive control may therefore be an important factor in the response to motor rehabilitation, and its causal role in response to rehabilitation should be examined.

## Data Availability

All data produced in the present study are available upon reasonable request to the authors

## Acknowledgments

We thank Dr. Oren Barzel and Dr. Adva Ressel Zviely for their clinical assistance. We thank the reviewers for their constructive comments. This work was supported by the Israeli Science Foundation grant 1244/22 to LS.

## Bibliography

Aprile, I., Guardati, G., Cipollini, V., Papadopoulou, D., Monteleone, S., Redolfi, A., Garattini, R., Sacella, G., Noro, F., Galeri, S., Carrozza, M. C., & Germanotta, M. (2021). Influence of Cognitive Impairment on the Recovery of Subjects with Subacute Stroke Undergoing Upper Limb Robotic Rehabilitation. Brain Sci, 11(5). 10.3390/brainsci11050587

Arbiv, D. C., & Meiran, N. (2015). Performance on the antisaccade task predicts dropout from cognitive training. Intelligence, 49, 25–31. 10.1016/j.intell.2014.11.009

Ballard, C., Stephens, S., Kenny, R. A., Kalaria, R., Tovee, M., & O’Brien, J. (2003). Profile of neuropsychological deficits in older stroke survivors without dementia. Dementia and Geriatric Cognitive Disorders, 16(1), 52–56. 10.1159/000069994

Bergman-Nutley, S., & Klingberg, T. (2014). Effect of working memory training on working memory, arithmetic and following instructions. Psychological Research, 78, 869–877. 10.1007/s00426-014-0614-0

Binyamin-Netser, R., Goldhamer, N., Avni, I., Ressel Zviely, A., & Shmuelof, L. (2023). Cognitive Impairments After Stroke Do Not Attenuate Explicit Visuomotor Adaptation in Reaching and Savings With the Unaffected Arm. Neurorehabilitation and Neural Repair, 37(7). 10.1177/15459683231177605

Boisgontier, M. P., Beets, I. A. M., Duysens, J., Nieuwboer, A., Krampe, R. T., & Swinnen, S. P. (2013). Age-related differences in attentional cost associated with postural dual tasks: Increased recruitment of generic cognitive resources in older adults. In Neuroscience and Biobehavioral Reviews (Vol. 37, Issue 8, pp. 1824–1837). 10.1016/j.neubiorev.2013.07.014

Brodtmann, A., Khlif, M. S., Egorova, N., Veldsman, M., Bird, L. J., & Werden, E. (2020). Dynamic Regional Brain Atrophy Rates in the First Year after Ischemic Stroke. Stroke, 183–192. 10.1161/STROKEAHA.120.030256

Chiti, G., & Pantoni, L. (2014). Use of Montreal Cognitive Assessment in Patients With Stroke. Stroke, 45(10), 3135–3140. 10.1161/STROKEAHA

Chow, W. Z., Ong, L. K., Kluge, M. G., Gyawali, P., Walker, F. R., & Nilsson, M. (2020). Similar cognitive deficits in mice and humans in the chronic phase post-stroke identified using the touchscreen-based paired-associate learning task. Scientific Reports, 10(1). 10.1038/s41598-020-76560-x

Cole, M. W., Bagic, A., Kass, R., & Schneider, W. (2010). Prefrontal Dynamics Underlying Rapid Instructed Task Learning Reverse with Practice. 10.1523/JNEUROSCI.1662-10.2010

Cole, M. W., Laurent, P., & Stocco, A. (2013). Rapid instructed task learning: a new window into the human brain’s unique capacity for flexible cognitive control. Cogn Affect Behav Neurosci, 13(1), 1–22. 10.3758/s13415-012-0125-7

Cole, M. W., Patrick, L. M., Meiran, N., & Braver, T. S. (2018). A role for proactive control in rapid instructed task learning. Acta Psychologica, 184, 20–30. 10.1016/J.ACTPSY.2017.06.004

Cumming, T. B., Marshall, R. S., & Lazar, R. M. (2013). Stroke, cognitive deficits, and rehabilitation: Still an incomplete picture. In International Journal of Stroke (Vol. 8, Issue 1, pp. 38–45). 10.1111/j.1747-4949.2012.00972.x

Dubois, J., Galdi, P., Paul, L. K., & Adolphs, R. (2018). A distributed brain network predicts general intelligence from resting-state human neuroimaging data. Philosophical Transactions of the Royal Society B: Biological Sciences, 373(1756). 10.1098/rstb.2017.0284

Dunham, S., Lee, E., & Persky, A. M. (2020). The Psychology of Following Instructions and Its Implications. 10.5688/ajpe7779

Finkel, D., Reynolds, C. A., McArdle, J. J., & Pedersen, N. L. (2005). The longitudinal relationship between processing speed and cognitive ability: Genetic and environmental influences. Behavior Genetics, 35(5), 535–549. 10.1007/s10519-005-3281-5

Friedman, N. P., & Miyake, A. (2004). The Relations Among Inhibition and Interference Control Functions: A Latent-Variable Analysis. Journal of Experimental Psychology: General, 133(1), 101–135. 10.1037/0096-3445.133.1.101

Goh, J. O., & Park, D. C. (2009). Neuroplasticity and cognitive aging: The scaffolding theory of aging and cognition. In Restorative Neurology and Neuroscience (Vol. 27, Issue 5, pp. 391–403). 10.3233/RNN-2009-0493

Graham, N. S. N., & Sharp, D. J. (2019). Understanding neurodegeneration after traumatic brain injury: from mechanisms to clinical trials in dementia. Journal of Neurology, Neurosurgery & Psychiatry, 90(11), 1221–1233. 10.1136/JNNP-2017-317557

Hartstra, E., Kühn, S., Verguts, T., & Brass, M. (2011). The implementation of verbal instructions: An fMRI study. Human Brain Mapping, 32(11), 1811. 10.1002/HBM.21152

Heuninckx, S., Wenderoth, N., & Swinnen, S. P. (2008). Systems neuroplasticity in the aging brain: Recruiting additional neural resources for successful motor performance in elderly persons. Journal of Neuroscience, 28(1), 91–99. 10.1523/JNEUROSCI.3300-07.2008

Hom, J., & Reitan, R. M. (1990). Generalized cognitive function after stroke. Journal of Clinical and Experimental Neuropsychology, 12(5), 644–654. 10.1080/01688639008401008

Hull, R., Martin, R. C., Beier, M. E., Lane, D., & Hamilton, A. C. (2008). Executive function in older adults: a structural equation modeling approach. Neuropsychology, 22(4), 508–522. 10.1037/0894-4105.22.4.508

Kase, C. S., Wolf, P. A., Kelly-Hayes, M., Kannel, W. B., Beiser, A., & D’Agostino, R. B. (1998). Intellectual decline after stroke: the Framingham Study. Stroke, 29(4), 805–812. 10.1161/01.str.29.4.805

Katzir, M., Eyal, T., Meiran, N., & Kessler, Y. (2010). Imagined positive emotions and inhibitory control: the differentiated effect of pride versus happiness. J Exp Psychol Learn Mem Cogn, 36(5), 1314–1320. 10.1037/a0020120

Laakso, H. M., Hietanen, M., Melkas, S., Sibolt, G., Curtze, S., Virta, M., Ylikoski, R., Pohjasvaara, T., Kaste, M., Erkinjuntti, T., & Jokinen, H. (2019). Executive function subdomains are associated with post-stroke functional outcome and permanent institutionalization. European Journal of Neurology, 26(3), 546–552. 10.1111/ene.13854

Levine, D. A., Galecki, A. T., Langa, K. M., Unverzagt, F. W., Kabeto, M. U., Giordani, B., & Wadley, V. G. (2015). Trajectory of Cognitive Decline after Incident Stroke HHS Public Access Exposure-Time-dependent incident stroke. JAMA, 314(1), 41–51. 10.1001/jama.2015.6968

McDowd, J. M., Filion, D. L., Pohl, P. S., Richards, L. G., & Stiers, W. (2003). Attentional abilities and functional outcomes following stroke. J Gerontol B Psychol Sci Soc Sci, 58(1), P45–53. 10.1093/geronb/58.1.p45

Meiran, N., Pereg, M., Givon, E., Danieli, G., & Shahar, N. (2016). The role of working memory in rapid instructed task learning and intention-based reflexivity: An individual differences examination. Neuropsychologia, 90, 180–189. 10.1016/j.neuropsychologia.2016.06.037

Meiran, N., Pereg, M., Kessler, Y., Cole, M. W., & Braver, T. S. (2015). The power of instructions: Proactive configuration of stimulus-response translation. J Exp Psychol Learn Mem Cogn, 41(3), 768–786. 10.1037/xlm0000063

Miyake, A., Emerson, M. J., & Friedman, N. P. (2000). Assessment of executive functions in clinical settings: problems and recommendations. Semin Speech Lang, 21(2), 169–183. 10.1055/s-2000-7563

Mok, V. C. T., Wong, A., Lam, W. W. M., Fan, Y. H., Tang, W. K., Kwok, T., Hui, A. C. F., & Wong, K. S. (2004). Cognitive impairment and functional outcome after stroke associated with small vessel disease. *Journal of Neurology*, Neurosurgery and Psychiatry, 75(4), 560–566. 10.1136/jnnp.2003.015107

Nasreddine, Z. S., Phillips, N. A., Bedirian, V., Charbonneau, S., Whitehead, V., Collin, I., Cummings, J. L., & Chertkow, H. (2005). The Montreal Cognitive Assessment, MoCA: a brief screening tool for mild cognitive impairment. J Am Geriatr Soc, 53(4), 695–699. 10.1111/j.1532-5415.2005.53221.x

Nys, G. M. S., van Zandvoort, ; M J E, de Kort, ; P L M, van der Worp, ; H B, Jansen, ; B P W, Algra, ; A, de Haan, ; E H F, & Kappelle, L. J. (2005). The prognostic value of domain-specific cognitive abilities in acute first-ever stroke. Neurology, 64(5), 821–827. www.neurology.org

Pereg, M., & Meiran, N. (2019). Rapid instructed task learning (but not automatic effects of instructions) is influenced by working memory load. PLoS ONE, 14(6). 10.1371/journal.pone.0217681

Peterburs, J., Pergola, G., Koch, B., Schwarz, M., & Hoffmann, K.-P. (2011). Altered Error Processing following Vascular Thalamic Damage: Evidence from an Antisaccade Task. PLoS ONE, 6(6), 21517. 10.1371/journal.pone.0021517

Pohjasvaara, T., Leskela, M., Vataja, R., Kalska, H., Ylikoski, R., Hietanen, M., Leppavuori, A., Kaste, M., & Erkinjuntti, T. (2002). Post-stroke depression, executive dysfunction and functional outcome. Eur J Neurol, 9(3), 269–275. https://www.ncbi.nlm.nih.gov/pubmed/11985635

Possin, K. L., LaMarre, A. K., Wood, K. A., Mungas, D. M., & Kramer, J. H. (2014). Ecological validity and neuroanatomical correlates of the NIH EXAMINER executive composite score. J Int Neuropsychol Soc, 20(1), 20–28. 10.1017/S1355617713000611

Rey-Mermet, A., Gade, M., & Oberauer, K. (2018). Should we stop thinking about inhibition? Searching for individual and age differences in inhibition ability. Journal of Experimental Psychology: Learning Memory and Cognition, 44(4), 501–526. 10.1037/XLM0000450

Roberts, R. J., Hager, L. D., & Heron, C. (1994). Prefrontal cognitive processes: Working memory and inhibition in the antisaccade task. Journal of Experimental Psychology: General, 123(4), 374–393. 10.1037/0096-3445.123.4.374

Roebers, C. M., & Feurer, E. (2016). Linking Executive Functions and Procedural Metacognition. Child Development Perspectives, 10(1), 39–44. 10.1111/CDEP.12159

Ruge, H., & Wolfensteller, U. (2010). Rapid Formation of Pragmatic Rule Representations in the Human Brain during Instruction-Based Learning. Cerebral Cortex, 20(7), 1656– 1667. 10.1093/CERCOR/BHP228

Salthouse, T. A. (1994). The nature of the influence of speed on adult age differences in cognition. Developmental Psychology, 30(2).

Salthouse, T. A. (2005). Relations between cognitive abilities and measures of executive functioning. Neuropsychology, 19(4), 532–545. 10.1037/0894-4105.19.4.532

Skidmore, E. R., Eskes, G., & Brodtmann, A. (2023). Executive Function Poststroke: Concepts, Recovery, and Interventions. In Stroke (Vol. 54, Issue 1, pp. 20–29). Wolters Kluwer Health. 10.1161/STROKEAHA.122.037946

Stuss, D. T., & Levine, B. (2002). Adult clinical neuropsychology: lessons from studies of the frontal lobes. Annu Rev Psychol, 53, 401–433. 10.1146/annurev.psych.53.100901.135220

Su, C. Y., Wuang, Y. P., Lin, Y. H., & Su, J. H. (2015). The role of processing speed in post-stroke cognitive dysfunction. Archives of Clinical Neuropsychology, 30(2), 148–160. 10.1093/arclin/acu057

Tatemichi, T. K., Desmond, D. W., Stern, Y., Paik, M., Sano, M., & Bagiella, E. (1994). Cognitive impairment after stroke: frequency, patterns, and relationship to functional abilities. J Neurol Neurosurg Psychiatry, 57(2), 202–207. 10.1136/jnnp.57.2.202

Verrico, C. D., Liu, S., Asafu-Adjei, J. K., Sampson, A. R., Bradberry, C. W., & Lewis, D. A. (2011). Acquisition and baseline performance of working memory tasks by adolescent rhesus monkeys. Brain Research, 1378, 91–104. 10.1016/j.brainres.2010.12.081

Wall, K. J., Isaacs, M. L., Copland, D. A., & Cumming, T. B. (2015). Assessing cognition after stroke. Who misses out? A systematic review. In International Journal of Stroke (Vol. 10, Issue 5, pp. 665–671). Blackwell Publishing Ltd. 10.1111/ijs.12506

Zinn, S., Bosworth, H. B., Hoenig, H. M., & Swartzwelder, H. S. (2007). Executive function deficits in acute stroke. Arch Phys Med Rehabil, 88(2), 173–180. 10.1016/j.apmr.2006.11.015

